# Arresting Vertical Transmission of Hepatitis B Virus (AVERT-HBV) in the Democratic Republic of the Congo

**DOI:** 10.1101/2021.03.19.21253929

**Authors:** Peyton Thompson, Camille E. Morgan, Patrick Ngimbi, Kashamuka Mwandagalirwa, Noro Ravelomanana, Martine Tabala, Malongo Fathy, Bienvenu Kawende, Jérémie Muwonga, Pacifique Misingi, Charles Mbendi, Christophe Luhata, Ravi Jhaveri, Gavin Cloherty, Didine Kaba, Marcel Yotebieng, Jonathan B. Parr

**Author notes:** **Corresponding Author:** Peyton Thompson, MD, MSCR; CB #7036, University of North Carolina School of Medicine, Chapel Hill, NC 27599. Co-second authors (Camille E. Morgan, Patrick Ngimbi).

## Abstract

**Background:** Hepatitis B virus (HBV) remains endemic throughout sub-Saharan Africa despite the widespread availability of effective vaccines. We evaluated the feasibility of adding HBV testing and treatment of pregnant women and birth-dose vaccination of HBV-exposed infants to the HIV prevention of MTCT (PMTCT) program infrastructure in the Democratic Republic of the Congo (DRC), where HBV treatment and birth-dose vaccination programs are not established.

**Methods:** As part of the HIV PMTCT program at two maternity centers in Kinshasa, DRC, pregnant women were screened for HBV at routine prenatal care registration. Pregnant women with high viral load and/or HBeAg positivity were offered tenofovir disoproxil fumarate (TDF). HBV-exposed infants received a birth-dose of HBV vaccine within 24 hours of life. The primary endpoint was the feasibility and acceptability of the study.

**Findings:** Of 4,016 women screened, 109 (2.7%) were HBsAg-positive. Ten of 91 (11.1%) women evaluated had high-risk disease. Of 88 infants, 60 (68.2%) received a birth-dose vaccine; of these, 46 (76.7%) received a timely birth-dose. No cases of HBV MTCT were observed in our cohort. There were no serious adverse events associated with TDF nor with birth-dose vaccine. The study procedures were highly acceptable (>80%) among mothers.

**Interpretation:** Adding HBV screening and treatment of pregnant women and infant birth-dose vaccination to existing HIV PMTCT platforms is feasible in countries like the DRC. Birth-dose vaccination against HBV integrated within the current Expanded Programme on Immunization (EPI) and HIV PMTCT program could accelerate progress toward HBV elimination in Africa.

## INTRODUCTION

Viral hepatitis is a leading infectious killer globally, with hepatitis B virus (HBV) alone accounting for nearly one million deaths annually.^1^ Despite the existence of an effective vaccine, the prevalence of HBV in sub-Saharan Africa (SSA) remains unacceptably high. Mother-to-child transmission (MTCT) is a silent driver of the HBV epidemic. In the absence of any intervention, 70% to 90% of mothers with high-risk HBV (defined as a viral load ≥ 200,000 IU/mL and/or HBV “e” antigen [HBeAg] positivity) will transmit the virus to their infants, compared to 10% to 40% of mothers who are HBeAg-negative.^2^ The World Health Organization (WHO) recommends that all infants receive a birth-dose of HBV vaccine as soon as possible after birth, preferably within 24 hours, followed by at least two additional vaccine doses.^3^ The three-dose series provides ≥95% protection against HBV infection, but breakthrough transmission can occur despite vaccination in infants born to women with high-risk HBV.^4,5^ In its 2020 guidelines on HBV prevention of MTCT (PMTCT), the WHO recommends that HBV-infected women with high-risk HBV receive antiviral prophylaxis from the 28^th^ week of pregnancy until or beyond delivery to prevent breakthrough MTCT.^3,6,7^ Most HIV-HBV co-infected women receive tenofovir as part of their antiretroviral regimen. However, the feasibility of providing antiviral prophylaxis for HBV mono-infected pregnant women in low-resource settings in SSA has yet to be tested.

The prevalence of HBV in the Democratic Republic of the Congo (DRC) is 3·3%, with high (2·2%) prevalence among children under age five, which correlates to approximately 450,000 Congolese children living with chronic HBV, and at risk for liver disease and premature death.^8,9^ While HBV vaccine has been part of the DRC’s Expanded Program on Immunization (EPI) since 2007,^10^ the first dose is not administered until six weeks of life, which is too late for the prevention of peri-partum MTCT. Similarly, there are no screening programs to identify and treat pregnant women with high-risk HBV, despite established infrastructure for the HIV PMTCT. The EPI delivers other birth-dose vaccines (BCG and polio), and so can be leveraged alongside the HIV PMTCT infrastructure to both deliver the birth-dose HBV vaccine and to screen and treat women with high-risk HBV.^11,12^

In this study, we assessed the feasibility of building upon existing EPI and HIV PMTCT infrastructures in Kinshasa, DRC to deliver HBV birth-dose vaccine and to identify and treat HBV-infected pregnant women at high risk of MTCT according to WHO guidelines.

## MATERIALS AND METHODS

### Study Setting and Design

This demonstration project was implemented in the two largest maternity centers in Kinshasa: Binza and Kingasani (both private/not-for-profit clinics). The two facilities were chosen not only because of their size, but also because of their long-standing collaborations with our HIV PMTCT team. In collaboration with the Expanded Program on Immunization (EPI), the required monovalent vaccine doses were acquired and stored within the existing cold chain system. We leveraged the infrastructure of an ongoing randomized trial of continuous quality interventions on long-term ART outcomes among pregnant/postpartum women in Kinshasa (CQI-PMTCT study, NCT03048669), through which we were already screening HIV-infected pregnant women for Hepatis B and C.^13,14^

### Study Population and Procedures

Between September 24, 2018 and February 22, 2019, all pregnant women registering for pretenatal services at either of the two clinics were provided group counseling on HBV and PMTCT thereof, and were invited to be screened for HBV. Those who verbally agreed were screened using point-of-care ALERE DETERMINE HBsAg testing (Abbott Diagnostics, Abbott Park, Illinois). Pregnant women who tested positive were referred for evaluation and inclusion in this feasibility study. To be eligible for prospective follow-up, pregnant women who screened HbsAg-positive needed to meet the following additional criteria: 1) gestational age ≤ 24 weeks (allowing time for viral load and HbeAg testing and return of results prior to initiation of tenofovir treatment between 28-32 weeks’ gestation), 2) intention to continue care at one of two maternity centers throughout the pregnancy and postpartum period, and 3) not sick enough to be hospitalized. Written informed consent was obtained in the participant’s preferred language (French or Lingala). At the time of the study, underage pregnant women (<18 years) were considered emancipated minors in the DRC and were allowed to consent directly without parental consent.

At the time of consent, a structured questionnaire was administered to collect information on demographic and clinical characteristics. All women were followed through 24 weeks postpartum, when they completed an exit questionnaire that assessed the acceptability of study procedures.

All participants underwent venous blood collection at baseline to determine risk status for HBV, including HBeAg (Abbott ARCHITECT HBeAg assay, Abbott Park, Illinois) and HBV DNA (Abbott RealTi*m*e HBV viral load assay, Abbott Park, Illinois), performed at the National Blood Transfusion Center and National AIDS Control Program laboratories in Kinshasa, respectively. Baseline kidney (blood urea nitrogen and creatinine) and liver (aspartate aminotransferase or AST, and alanine aminotransferase or ALT) function was evaluated in all enrolled women. Women with a viral load ≥ 200,000 IU/mL and/or HBeAg positivity were considered at ‘high-risk’ for HBV MTCT and were further assessed for eligibility for prophylactic treatment with tenofovir disoproxil fumarate (TDF).^3,15,16^ Women with high-risk HBV, without signs of reduced kidney function (glomerular filtration rate (GFR) <50 mL/min) or abnormal liver function (as determined by the study physician), were initiated on once daily 300mg dosing of TDF (Gilead Sciences, South San Francisco, CA) treatment between 28-32 weeks’ gestation and continued through 12-weeks’ postpartum. We assured that any women with HIV-HBV co-infection were on a TDF-containing regimen, per national guidelines. Details of additional follow-up procedures for women with high-risk HBV can be found in the **Supplementary Material**. To assess TDF adherence, tenofovir diphosphate (TFVdp) levels were measured at the University of North Carolina’s Eshelman School of Pharmacy using DBS samples collected from high-risk women at delivery. TFVdp levels represent an average of expected adherence over a several month period, as the medication has a long half-life in red blood cells. TFVdp concentrations in DBS of 490, 980 and 1750 fmol/punch indicated a participant taking 2, 4 and 7 doses per week, respectively, based on an expected 1.4-fold bias in concentrations from published thresholds.^17^

All exposed infants were given a birth-dose of monovalent HBV vaccine (Euvax-B, 0·5 mL), as soon as possible and ideally within 24 hours of life. The timing of birth and of vaccination were recorded. Infants were monitored for at least 24 hours after receipt of vaccine, and adverse reactions were recorded. Infants underwent point-of-care HBsAg testing at 24 weeks.

### Measures and Statistical Analysis

Feasibility was the key part of the program that was evaluated: the feasibility of screening pregnant women to identify those at high risk for HBV MTCT and to provide them with antiviral prophylaxis, and the feasibility of administering the birth-dose vaccine to infants of women with HBV.

To assess the feasibility of viral prophylaxis for women at high risk of HBV MTCT, we calculated the following proportions: 1) women who tested positive by HbsAg screening, 2) HBsAg+ women who consented for enrollment, 3) eligible women with high-risk HBV who accepted TDF prophylaxis, and 4) women on TDF who adhered to treatment as measured by a) pill count, b) viral suppression at delivery (defined as HBV viral load < 200,000 IU/mL), and c) TFVdp level. Acceptability of the program was assessed by calculating frequencies of aggregate responses on the exit questionnaire. A predetermined cutoff of 80% reporting the program to be “very acceptable” or “somewhat acceptable” on survey responses was deemed “acceptable”.

Secondarily, we evaluated the safety of TDF prophylaxis by screening for adverse events, and the percentage of women who developed ALT elevation during/after antiviral treatment. Timeliness of birth-dose vaccination was calculated as the proportion of infants who received birth-dose vaccine within 24 hours of life. The incidence of HBV MTCT (the proportion of infants who tested positive for HBsAg at 24 weeks) was calculated. Finally, we calculated the proportion of women who were lost-to-follow-up (LTFU) by delivery and by 24 weeks’ postpartum.

Demographics of enrolled women were analyzed from the enrollment questionnaire. A principal component analysis was used to combine household attributes into a wealth index similar to other Kinshasa-based studies (**Supplementary Material**).^18^ Statistical computations were carried out using R 4.0.3 (R Core Team, Vienna, Austria).

Institutional Review Board approval was obtained from the University of North Carolina at Chapel Hill (#17-2090) and the Kinshasa School of Public Health (CE/ESP/001/2018) prior to initiation of the study. The trial is registered at ClinicalTrials.gov (NCT03567382).

## RESULTS

### Maternal HBV prevalence and high-risk disease

Of 4,016 pregnant women screened for HBV, 109 were HBsAg-positive, indicating an overall HBsAg prevalence of 2·7% (95% CI: 2·2%-3·2%). A total of 91 women met eligibility criteria; all provided informed consent and were enrolled. Enrollment and results across the care continuum are displayed in **Figure 1**. One woman was withdrawn from the treatment program due to a false pregnancy (study staff confirmed a negative urine pregnancy test despite report of pregnancy by the woman); data from 90 women were therefore included. The median age of enrolled women was 31 (IQR: 25,34) years and the median gestational age was 19 weeks (IQR: 15,22) (**Table 1**). Only one woman was co-infected with HIV and HBV; she received a TDF-based antiretroviral regimen. Reasons women elected not to enroll are shown in **Figure 2**. Of the 90 women, ten (11·1%) had high-risk HBV; nine of ten were HBeAg positive and five of ten had a high viral load. Women with high-risk HBV were younger and presented later in pregnancy than women without high-risk HBV (**Table 1**).

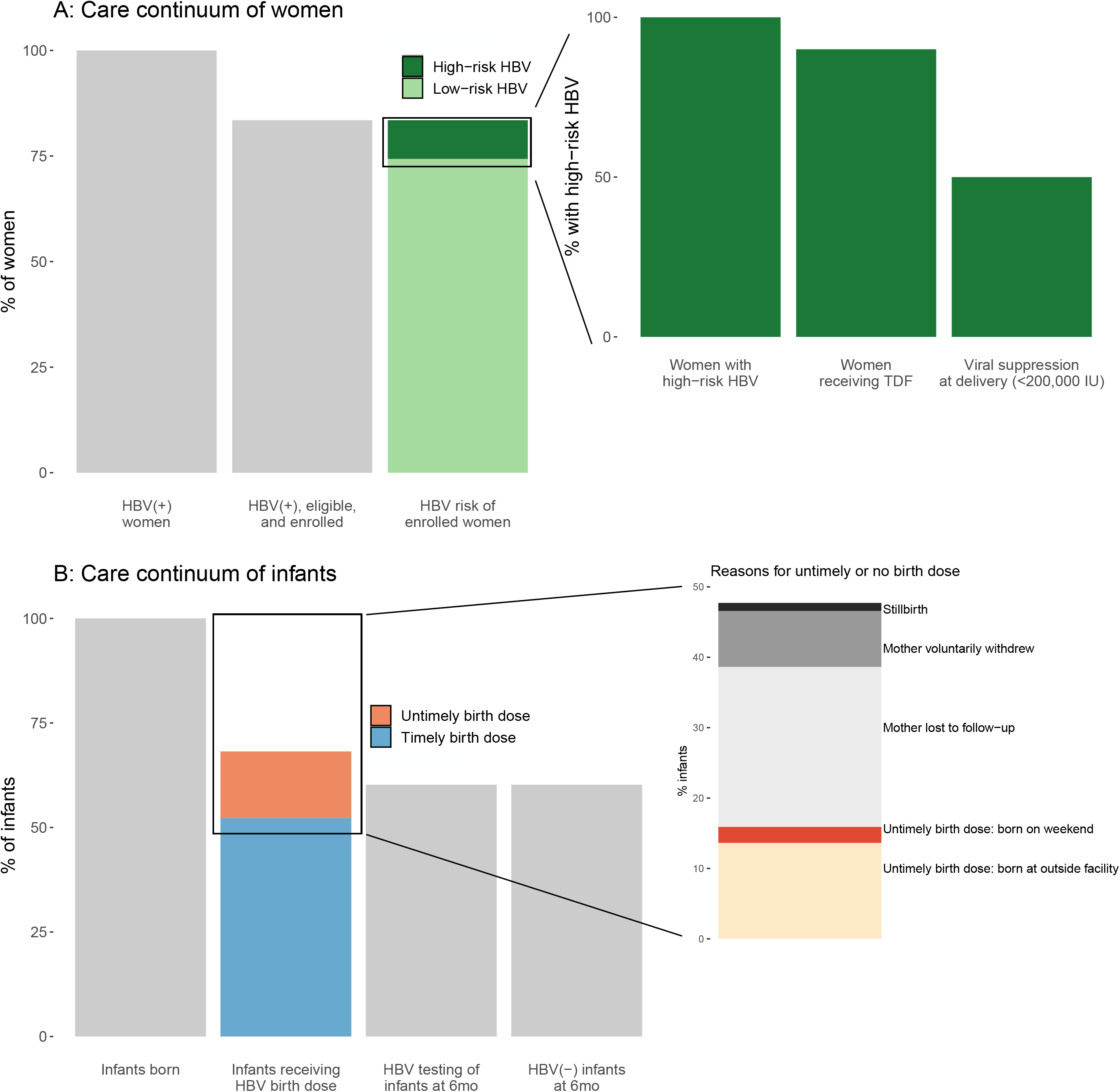

**Table 1.**
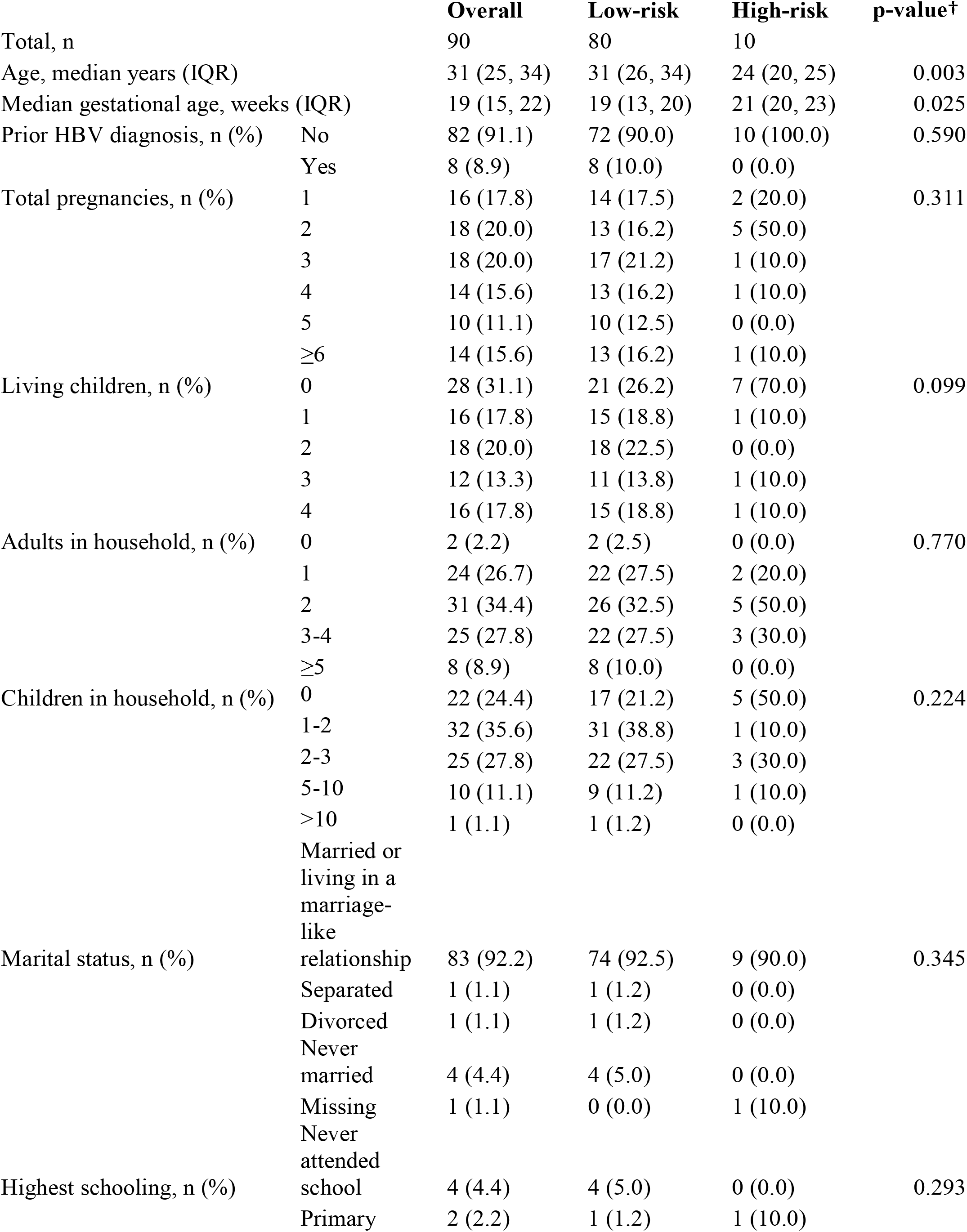
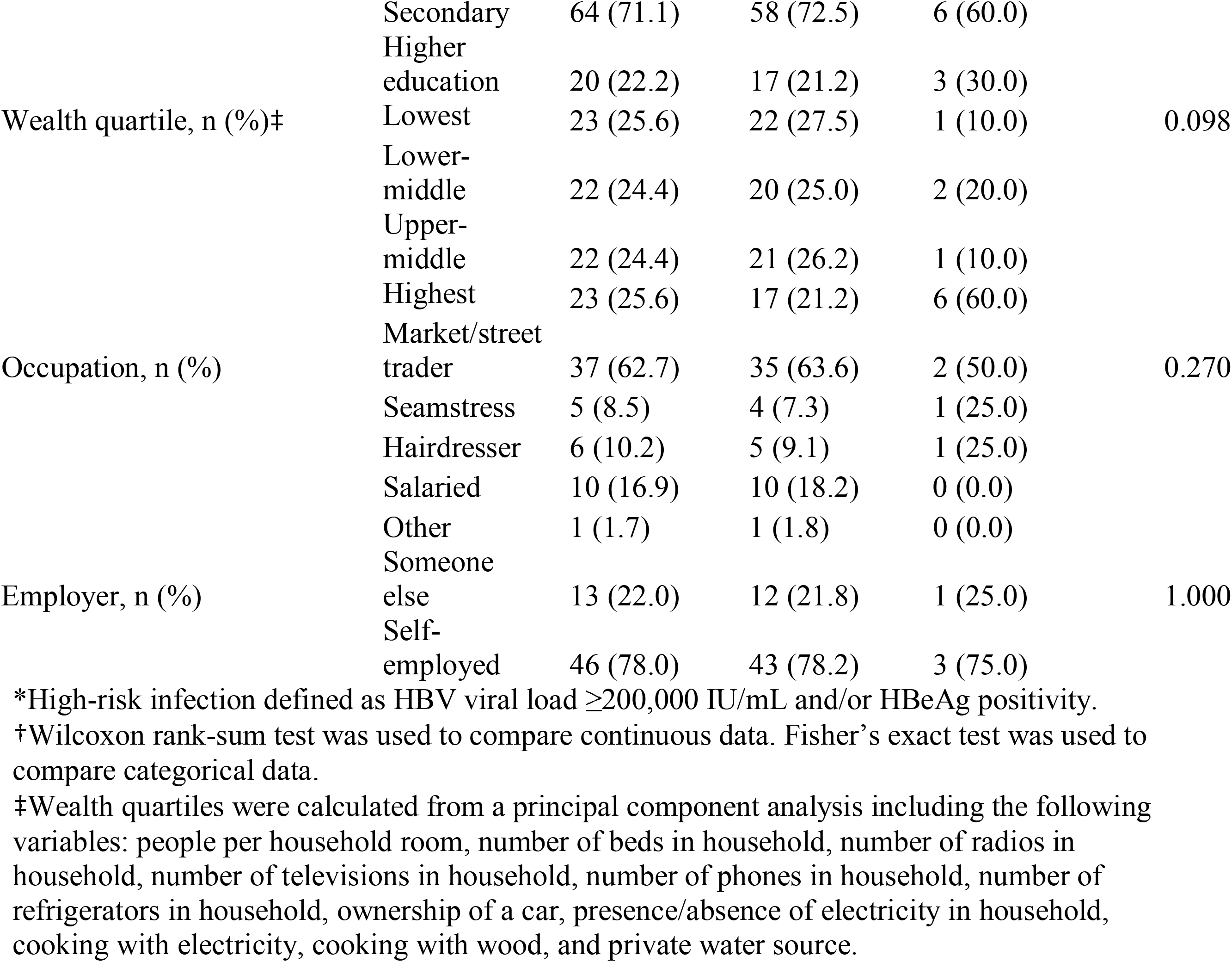
Characteristics of the study population, by high- and low-risk HBV infection*.

**Figure 2.**
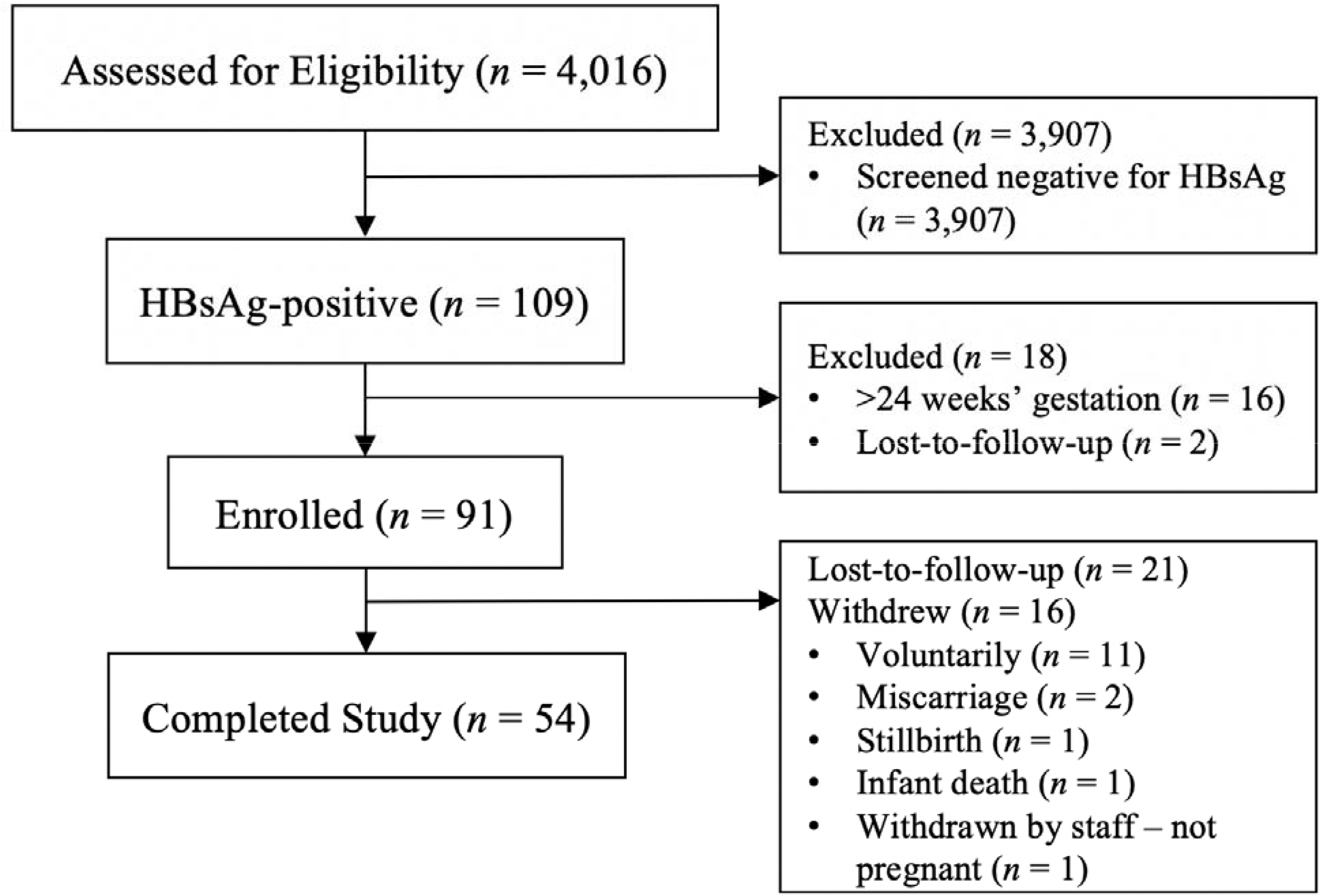
Flowsheet of screened women and reasons for not enrolling.

### Retention in care through delivery and 24-weeks’ postpartum

Three women voluntarily withdrew and 12 were lost-to-follow-up by the time of the delivery visit. Eleven women voluntarily withdrew by 24-weeks’ postpartum, three women did not have living children (two miscarriages and one stillbirth), one infant died, and 21 women were lost-to-follow-up despite repeated efforts by the study team to contact them by telephone or in person (**Figure 2**). Thus, 54 women (60·0%) completed the study through the 24-week postpartum visit. Of the ten women with high-risk HBV, six (60·0%) were followed to study completion. Two women with high-risk HBV voluntarily withdrew from the study; the first prior to initiation of TDF, and the second after delivery. A third woman moved to another province in the DRC after delivery, and a fourth was lost to follow-up (LTFU) after the 10-week visit.

### Infant vaccination and MTCT

A total of 88 infants were born to enrolled women. Overall, sixty of 88 infants (68·2%) received a birth-dose of monovalent HBV vaccine; of these, 46 (76·7%) received a timely birth-dose within 24 hours. Among 63 infants born at the two study facilities, 48 (76·2%) received a birth-dose, and 41 of 48 (85·4%) were timely. Among 25 infants born at an outside facility, 12 (48·0%) received a birth-dose, and five of 12 (41·7%) were timely. None of the infants were born outside of a health facility. **Figure 1B** depicts the care continuum for infants, including reasons for missed or untimely vaccination. No adverse events were reported related to the birth-dose. Of 53 infants tested at 24 weeks, zero (0%) tested positive for HBsAg, consistent with no HBV MTCT.

### TDF adherence and viral suppression

Nine of ten (90·0%) women with high-risk HBV were initiated on TDF. One woman refused therapy and withdrew from the study. Five of nine (55·6%) women achieved viral suppression (<200,000 IU/mL) on TDF therapy by the time of delivery (**Figure 1A**); the remaining four women had decreased viral loads from enrollment to delivery, but not below 200,000 IU/mL (**Figure 3**). While all women taking TDF returned empty pill bottles and reported full adherence per the exit survey, TFVdp levels suggested low adherence. Three of nine women delivered within 28 days of initiation of TDF therapy, so TFVdp cannot be interpreted. Of the remaining six, three women had levels below the limit of quantification (BLQ), and one each with levels reflecting <2 doses per week, 2-4 doses per week, and 4-7 doses per week (**Figure 3**). All three women with detectable TFVdp who initiated TDF therapy >28 days prior to delivery were virally suppressed at delivery (**Figure 3**).

**Figure 3.**
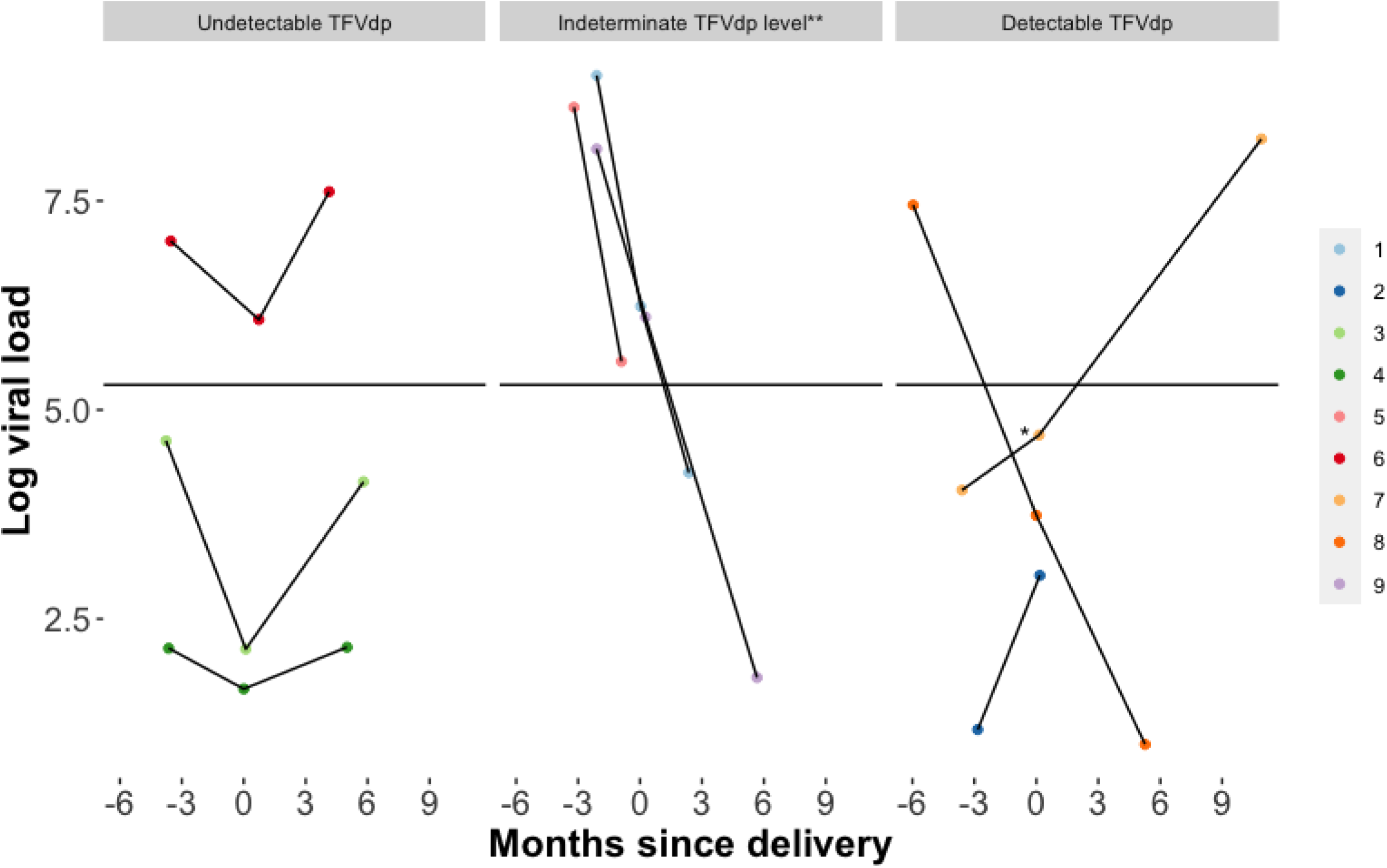
Viral loads of women with high-risk disease across study timepoints by tenofovir diphosphate (TFVdp) level. TFVdp levels reflect average adherence over several months. * Imputed viral load from dried blood spot assay (see Supplementary Materials). * * Tenofovir disoproxil fumarate initiated within 28 days of delivery, before TFVdp steady-state is expected.

### TDF safety

There were no serious adverse events associated with TDF. One woman reported dizziness while taking the medication; otherwise, no side effects were reported. Three women (33·3%) developed rebound elevations of ALT (**Supplementary Figure 1**), and two women (22·2%) developed corresponding elevations of HBV DNA after stopping TDF at 12-weeks’ postpartum. These women were referred to the study gastroenterologist for further management, but none attended their appointments.

### Acceptability of program procedures

All seven high-risk women followed through six months reported the act of taking TDF as “very acceptable”. Forty-eight of 54 women (88·9%) followed through 24-weeks’ postpartum reported having their blood drawn as “very acceptable,” with only two women reporting this as “very” or “somewhat unacceptable” (**Figure 4**). Only four women reported the infant blood draw as “somewhat” or “very unacceptable”. Only one woman (1·9%) reported vaccination of her infant as “very unacceptable”.

**Figure 4.**
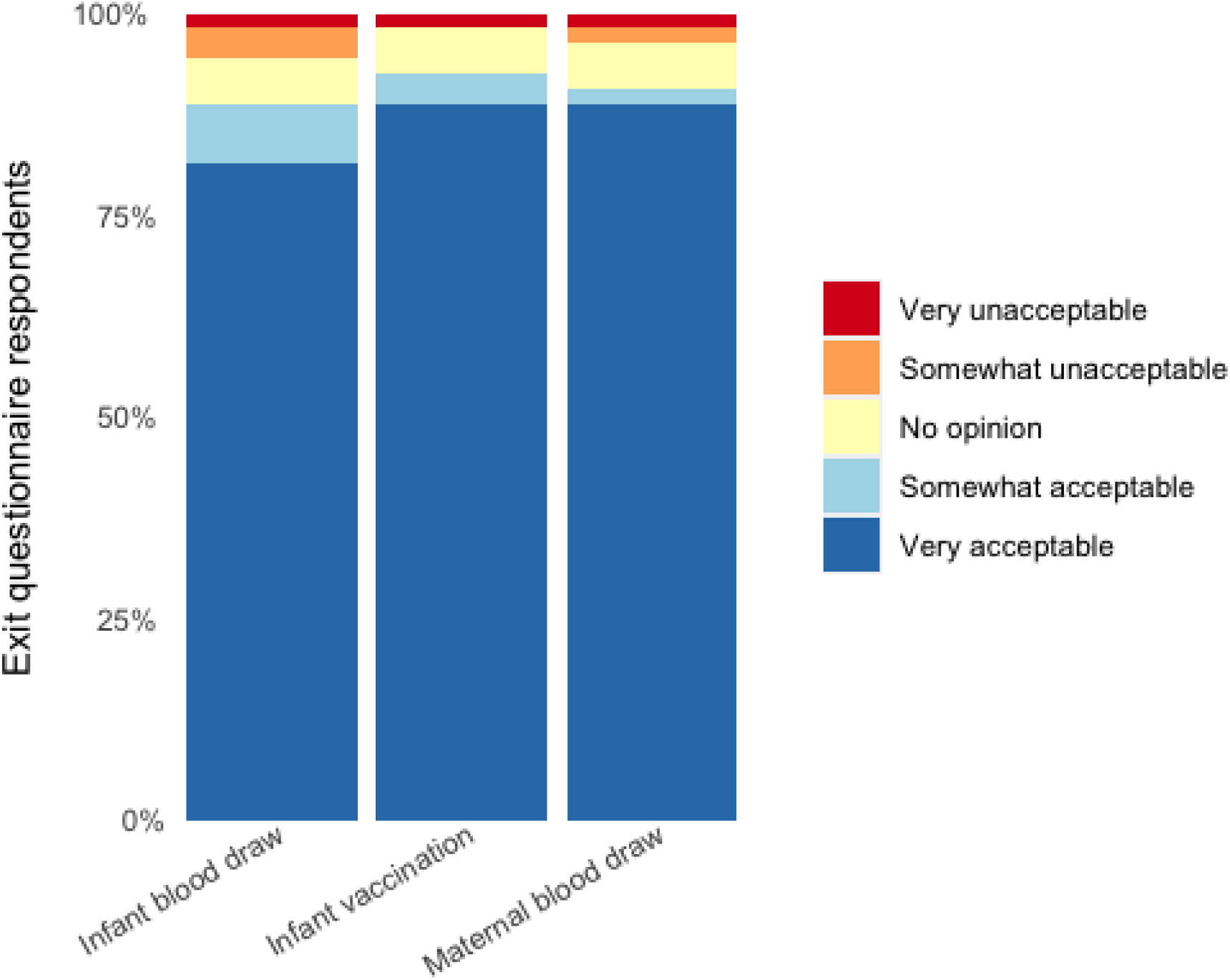
Acceptability of clinical study components assessed at 6-month follow-up visit (n=54)

## DISCUSSION

We demonstrate the feasibility of implementing a PMTCT of HBV program in the DRC using existing HIV PMTCT and EPI infrastructure with a two-pronged approach: 1) screening and treatment of HBV-positive pregnant women at high-risk for MTCT, and 2) provision of a birth-dose vaccine to HBV-exposed infants. Of the 53 HBV-exposed infants followed through 24 weeks, none were positive for HBV, indicating no vertical transmission. Even with vaccination, the estimated risk of MTCT among infants born to women with high-risk HBV ranges from 20% in Asia to 32% in Africa, compared to less than 1% for vaccinated infants born to Asian and African women without high-risk features.^19^ This evidence prompted the WHO to revise HBV PMTCT guidelines in 2020 to add antiviral prophylaxis to the recommended three-dose vaccination, with the first dose within 24 hours of life.^3^ Our successful integration of HBV screening of pregnant women and provision of TDF prophylaxis within an exisiting HIV PMTCT program in the two busiest maternity centers in Kinshasa is evidence to support the feasibility of this WHO recommendation, even in the most resource-limited settings.

The prevalence of HBV among pregnant women in this study was in line with similar studies in African settings.^19,20^ We found that 11·1% of HBV-positive women met criteria for TDF therapy based on high viral load and/or HBeAg positivity, a percentage that is similar to other reports in SSA.^20-22^ The assessment of high-risk HBV and administration of TDF was feasible within this study’s framework, but without reliable point-of-care tests for HBV DNA quantification or HbeAg testing, scale-up will be a challenge. Until such infrastructure is in place, HBV birth-dose should be prioritized as a prevention tool.^23-25^

Birth-dose vaccination was feasible in two high-volume maternity centers in urban Kinshasa, DRC, and was largely acceptable to mothers. While we did not achieve vaccination within 24 hours of birth for all infants, this feasibility study demonstrates that a higher standard-of-care is within reach in the DRC. We identified several barriers to timely administration of the birth-dose that must be overcome as the country moves to universal birth-dose vaccination. First, many women delivered at outside facilities rather than at one of the two study facilities. Potential reasons for delivering at outside facilities include personal preference, cost, quality of care received and/or need for C-section delivery (which was not possible at either maternity hospital in the present study). With the adoption of a universal HBV birth-dose vaccine, all facilities would have access such that maternal preference of delivery site would not determine whether the infant is vaccinated. Second, baseline knowledge of HBV was limited among mothers in our study (unpublished data), and dropouts along the care cascade were relatively high. Our team has previously shown that a mother’s perception of HIV risk to her infant was a key determinant of her retention in the HIV PMTCT care cascade.^26^ Third, reliable cold chains are needed. With support from Gavi and the DRC EPI, established cold chains for other routine vaccines could be strengthened and used for HBV birth-dose vaccine. At the same time, sentinel studies have already evaluated storage of HBV birth-dose vaccine outside of the cold chain.^27^ Finally, many births occur outside of health facilities in the DRC and throughout SSA. Initial roll-out to health centers could be followed by decentralized vaccination after home birth, although the logistics of this strategy are more complex. Nonetheless, with small investments of resources to strengthen the existing EPI infrastructure in the DRC, timely delivery of the HBV birth-dose is possible alongside other birth-dose vaccinations.^28^

We uncovered a discordance between verbally reported adherence to TDF therapy and adherence as measured by TFVdp levels. Only three of nine women had TFVdp levels suggesting that they took at least two doses per week; two of these three women had viral load suppression at delivery. This is in contrast to the nearly 100% adherence reported on exit surveys and via pill counts (only empty bottles were returned). Changes in viral load and creatinine among women taking TDF can be used as proxy measures of adherence but do not fully explain the discordance. Further studies are needed to understand the drivers of poor adherence.

The strength of this study lies in its pragmatic, comprehensive approach. In line with recent WHO guidelines for preventing HBV MTCT, we integrated HbsAg screening into an existing HIV PMTCT program, leveraged laboratory infrastructure to identify and treat pregnant women with high-risk HBV, and leveraged the existing EPI cold chain infrastructure to deliver birth-dose vaccination to exposed infants. This package of interventions is delivered routinely to prevent HBV MTCT in the Global North, but not in Africa. We longitudinally followed HBV-infected women and their exposed infants through 24 weeks’ postpartum to document adherence to the cascade of care, side effects of birth-dose vaccine and TDF, and incidence of MTCT. While most studies of HBV MTCT in Africa published to-date focus on HBV-HIV co-infection, our cohort of mainly HBV mono-infected pregnant women is generalizable to settings in Africa where HIV prevalence is low.

This study also had limitations. First, the sample size was relatively small. Yet, to the best of our knowledge, this is the largest study of PMTCT of HBV among mono-infected pregnant women in SSA performed to-date. Second, the absence of a control group prevented us from quantifying the effectiveness of specific interventions. We were ethically obligated to provide effective HBV prevention measures; other studies in African settings have described the natural history of chronic HBV.^29^ Third, we observed a high rate of LTFU in this study. While LTFU negatively impacts our ability to determine incidence of MTCT, it is an expected feature of pragmatic feasibility studies and consistent with postnatal follow-up patterns in the DRC.^18^ Fourth, supply chain disruption prevented us from completing planned laboratory comparisons of HBeAg and viral load testing on DBS versus venous blood in-country. The COVID-19 pandemic bears some responsibility for these challenges, but nascent HBV laboratory reagent supply chains are also to blame. Delays in the return of laboratory results to some participants may have contributed to withdrawal or LTFU. Unanticipated stockouts led to a lack of HBeAg results for three women at enrollment; all three women were classified as low-risk for HBV MTCT based on their low viral loads. Finally, the measurement of TFVdp levels from DBS may not accurately depict adherence to TDF therapy. TFVdp is not sensitive for behavior change, as it represents an average of adherence over several months. Furthermore, we cannot exclude the possibility of TFVdp degradation during DBS preparation or transportation from the DRC, where ambient temperatures and humidity are high. There is no clear threshold for how much tenofovir is required to prevent HBV MTCT, but decreases in maternal HBV DNA prior to delivery are associated with decreased risk of HBV MTCT.^6,7^ Despite variable TFVdp levels, most women did have a several log decline in HBV DNA. Even with these limitations, we successfully implemented effective HBV PMTCT measures.

In conclusion, this study provides strong evidence in support of the WHO recommendation that mother-to-infant HBV transmission can be prevented by building upon HIV PMTCT and EPI platforms in resource-constrained settings such as the DRC. Lessons learned from HIV PMTCT, including how to improve retention and adherence to prophylaxis, can be leveraged to address similar challenges in the HBV PMTCT program. Deploying a package of proven interventions used routinely in the Global North but not widely adopted in Africa, we demonstrate the feasibility of HBV MTCT prevention measures during routine care at two high-volume maternity hospitals in Kinshasa. A two-pronged maternal screening/treatment and infant birth-dose approach was acceptable to mothers, and no cases of HBV MTCT were observed. Though obstacles must be overcome before roll-out of antiviral prophylaxis for pregnant women at high-risk of HBV MTCT, universal birth-dose HBV vaccination is a proven measure that should be implemented in the DRC and throughout SSA. To meet WHO elimination goals by 2030,^30^ countries with endemic HBV such as the DRC should adopt and implement WHO guidelines that establish pathways for HBV PMTCT,^3^ and should prioritize cross-cutting HBV prevention measures that include birth-dose vaccination.

## Data Availability

Datasets will not be made publicly available because they contain protected health information. However, the authors will share de-identified participant data, alongside the study protocol, statistical analysis plan and analytic code, upon reasonable request and with approval by an independent review committee..

## Abbreviations

ALT: Alanine aminotransferase
AST: Aspartate aminotransferase
DBS: Dried Blood Spot
DRC: Democratic Republic of the Congo
HBeAg: Hepatitis B envelop Antigen
HBsAg: Hepatitis B surface Antigen
HBV: Hepatitis B Virus
MTCT: Mother-to-child Transmission
PMTCT: Prevention of Mother-to-child Transmission
SSA: Sub-Saharan Africa
TDF: Tenofovir Disoproxil Fumarate

## NOTES

### Contributors

PT, JBP, RJ, MY and SRM designed the study. PT wrote the first draft, with contributions from CEM, MY and JBP. KM, NR, BK, MF, MT, PN, PM and CM contributed to field work. CEM, PT, and JBP analyzed the data. All authors edited and revised the final manuscript and approve it in its final version.

## Acknowledgements

This project was supported by a Gillings Innovation Laboratory award funded by the 2007 Gillings Gift to UNC-Chapel Hill’s Gillings School of Global Public Health. Abbott Laboratories provided HBV reagents free-of-charge for this study. The HIV PMTCT study whose infrastructure was leveraged for this project was supported by a grant from the National Institute of Health (NICHD R01HD087993). TFVdp testing was performed by the UNC Center for AIDS Research Clinical Pharmacology and Analytical Chemistry Core, an NIH funded program (P30AI050410). PT is supported by a grant from the National Institute of Health (NIAID K08AI148607). CEM received support from a grant from the National Institute of Health (NIGMS T32GM008719).

We would like to thank all of the women who participated in this study. We also acknowledge the staff at the maternity clinics, as well as provincial and national health authorities. We are grateful for the support we have received from administrative staff at the University of North Carolina and at Kinshasa School of Public Health. We also thank David Wohl for helpful advice and and the UNC Clinical Pharmacology and Analytical Chemistry Core for supporting the TFVdp assays. We grieve the loss of Professor Steven Meshnick, who played a major role in this study and whose vision and mentorship were critical to its success.

## Data Sharing Statement

Datasets will not be made publicly available because they contain protected health information. However, the authors will share de-identified participant data, alongside the study protocol, statistical analysis plan and analytic code, upon reasonable request and with approval by an independent review committee (“learned intermediary”).

