## Supplemental Material for "Arresting Vertical Transmission of Hepatitis B Virus (AVERT-HBV) in the Democratic Republic of the Congo"

**SUPPLEMENTARY MATERIAL**

**SUPPLEMENTARY METHODS**

*Statistical analysis*

We used *prcomp()* in R.4.0.3 to conduct a principal component analysis to calculate wealth quartiles. While this is an imperfect measure of socioeconomic status (SES), this method has become more common and accepted given the challenges in assessing SES.^1,2^ The following variables were included and scaled (*scale=TRUE* in *prcomp()*): an inverted categorization of people per household room (since fewer people per room suggests less crowding, which is associated with poverty), number of beds in household, number of radios in household, number of televisions in household, number of phones in household, number of refrigerators in household, ownership of cars in household (yes/no), electricity in household (yes/no), cooking with electricity (yes/no), cooking with wood (yes/no), and private water source (yes/no). The first component explained 23.5% of the variance and following similar studies,^3^ was used to calculate wealth quartiles using 25^th^, 50^th^, and 75^th^ percentiles of this variable.

*DNA extraction from dried blood spots (DBS)*

Venous blood samples were used for clinical management, however DBS samples were collected for research purposes and used to impute a single missing venous blood HBV DNA value (Figure 2). DNA was extracted from a single 6-mm DBS punch using Chelex and Tween as previously described.^4^ To prevent DNA contamination during DBS punching, hole punchers were dipped into 100% ethanol, passed under a flame, cooled to ambient temperature, and used to punch blank Whatmann filter paper five times between each sample. HBV DNA was extracted using a two-day Tween-Chelex protocol adapted from Teyssier *et al*. 2019.^4^ Briefly, 1 mL of 0·5% Tween 20 in 1X PBS was placed in each sample well, which contained a single 6mm DBS punch. All extractions were performed with two wells containing a 6mm DBS punch blank as negative controls, and the plate was then incubated overnight at room temperature on a shaker. On day two, the plate was centrifuged for 1 minute at 300 rpm, and Tween-PBS was aspirated from each well. Each sample was washed with 1 mL of 1X PBS and incubated for 20 minutes at 4°C. Following repeat centrifuging and aspiration of the PBS, 150 µL of a 1:2 solution of 20% Chelex-100 resin in water was added to each well and the plate was incubated for 10 minutes at 95°C, vortexing every two minutes. The plate was then centrifuged for 10 minutes at 1500 rpm and the supernatant transferred to a 96-well plate, which was centrifuged for five minutes at 1500 rpm. The supernatant from this final step was transferred to an Eppendorf LoBind DNA 96-well plate for storage. Following quantitative PCR (qPCR; described below), the extracted DNA were stored at -80°C.

*DNA quantification*

Samples underwent qPCR, along with two negative controls (water) and serially diluted synthetic HBV DNA standards (VR-3232SD, ATCC, Manassas, VA), two each per concentration of viral load: 50,000, 5000, 500, and 50 copies/mL. Real time PCR was conducted in 20 µL volumes, using the AB Taqman Universal Master Mix and 4 µL of sample DNA. HBV-DNA primer and probe sequences and concentrations are summarized in **Supplementary Table 1**, adopted from Welzel *et al*, 2006.^5^ The PCR cycles were: 2 minutes at 50°C, 10 minutes at 95°C, and 45 cycles of 15 seconds of denaturation at 95°C and 1 minute of annealing at 60°C. Viral loads were calculated using IU/mL.

**Supplementary Table 1: HBV primers and probe used for qPCR**

| Description | Final concentration | Sequence (5’ to 3’) |
| --- | --- | --- |
| HBV-F3 (for) | 500 nM | GGCCATCAGCGCATGC |
| HBV-R3M3 (rev) | 500 nM | C [5-NitIdl] GCTGCGAGCAAAACA |
| HBV-P3 (probe) | 100 nM | FAM-CTCTGCCGATCCATACTGCGGAACTC-TAMRA |

*Imputation of missing venous blood viral load from DBS*

The low sample size of participants with venous blood and DBS HBV DNA qPCR values within the assays’ upper and lower limits of quantification of each assay was not conducive to a valid regression fit line for imputation of the venous blood viral load for ID 7 at delivery. As an alternative, we compared venous blood and DBS concentrations across all samples (**Supplementary Table 2**). Based on DBS qPCR results, we conservatively estimated a venous HBV viral load of 50,000 IU/mL for the single sample without venous PCR results (ID 7, see Figure 2).

**Supplementary Table 2: Venous blood and DBS viral loads by participant and timepoint**

| **Participant ID** | **Study timepoint** | **Venous blood viral load (IU/mL)** | **DBS viral load (IU/mL)*** |
| --- | --- | --- | --- |
| 8 | 6-mo follow-up | 10 | 0.1839435 |
| 2 | Enrollment | 15 | Undetermined |
| 4 | Delivery | 46 | Undetermined |
| 9 | 6-mo follow-up | 63 | Undetermined |
| 3 | Delivery | 138 | Undetermined |
| 4 | Enrollment | 141 | Undetermined |
| 4 | 6-mo follow-up | 145 | Undetermined |
| 2 | Delivery | 1047 | Undetermined |
| 10 | Enrollment | 2951 | Undetermined |
| 8 | Delivery | 5495 | 0.15656285 |
| 7 | Enrollment | 10,965 | 0.16548684 |
| 3 | 6-mo follow-up | 13,804 | Undetermined |
| 7 | **Delivery**** |  | 0.26327923 |
| 3 | Enrollment | 42,658 | 1.6400603 |
| 5 | Delivery | 380,189 | 11.816989 |
| 6 | Delivery | 1,202,264 | 169.953 |
| 9 | Delivery | 1,288,250 | 56.9374 |
| 1 | Delivery | 1,737,801 | 6.6057897 |
| 8 | Enrollment | 28,183,829 | 225.81763 |
| 6 | 6-mo follow-up | 40,738,028 | 234.611 |
| 9 | Enrollment | 131,825,674 | 1168.0933 |
| 7 | 6-mo follow-up | 173,780,083 | 1340.2036 |
| 5 | Enrollment | 416,869,383 | 638.2598 |
| 1 | Enrollment | >1,000,000,000 | 130.30763 |

*Values below 10 IU/mL should not be interpreted linearly.

**Sample of interest for imputation of venous blood viral load.

*Additional laboratory evaluation of women with high-risk HBV*

Women with high-risk HBV underwent platelet testing at enrollment, as well as additional liver function testing at delivery, 10 weeks’ and 24 weeks’ postpartum. Additional follow-up procedures included: 1) assessments for women on TDF at monthly visits and referral of those with symptoms and signs suggestive of potential side effects to the study physician, and 2) assessment of adherence to TDF prophylaxis – by pill count, verbal report and tenofovir levels. Women returned their pill bottles at each monthly visit and the number of remaining pills were counted to determine how many were consumed since the last visit. Women also reported adherence verbally at the time of the exit visit (24 weeks’ postpartum). Combining TFVdp data with viral load data and kidney and liver function testing, we observed that women with detectable TFVdp who began TDF ≥ 28 days prior to delivery (n=3) were virally suppressed at delivery and had an average creatinine change of +43%. Women who began TDF within 28 days of delivery (n=3) had decreasing viral loads across all timepoints, but not virally suppressed at delivery, and had an average creatinine change of -4·8%. Women without detectable TFVdp (n=3) had decreasing viral loads at delivery that subsequently increased at six months (two remained below 200,000 IU/mL), and had an average creatinine change of +3·3%. Mild increases in serum creatinine are common during TDF therapy, thus serving as an indirect measure of adherence. **(Supplementary Table 3).**

**Supplementary Table 3: Individual laboratory trends for high-risk women by TFVdp levels**

| **Participant ID** | **Detectable TFVdp** | **TDF initiation >28 days from delivery?** | **Viral load trend (viral suppression defined as <200,000 IU/mL)** | **Creatinine at enrollment to delivery (% change)** | **AST (upper row, U/L) and ALT (lower row, U/L)** | | | |
| --- | --- | --- | --- | --- | --- | --- | --- | --- |
|  |  |  |  |  | Enrollment | Delivery | 10 weeks | 6 months |
| 2 | Yes | Yes | Increase at delivery but still virally suppressed | 0.38 to 0.57 (+50.0%) | 12 | 23 | 16 | 28 |
|  |  |  |  |  | 15 | 19 | 19 | 19 |
| 7 | Yes | Yes | Increased at delivery but still virally suppressed | 0.67 to 0.80 (+19.4%) | 29 | 58 | - | **253*** |
|  |  |  |  |  | 25 | **89*** | - | **192*** |
| 8 | Yes | Yes | Reached and sustained viral suppression | 0.60 to 0.96 (+60.0%) | 22 | 20 | 33 | 13 |
|  |  |  |  |  | 16 | 40 | 38 | 15 |
| 5 | Yes | No | Decreased at delivery but not virally suppressed | 0.72 to 0.59  (-18.1%) | 18 | 29 | 22 | - |
|  |  |  |  |  | 20 | 50 | 21 | - |
| 9 | Yes | No | Decreased and virally suppressed by 6-months | 0.76 to 0.68  (-10.5%) | 15 | 28 | 22 | 19 |
|  |  |  |  |  | 15 | 27 | 18 | 15 |
| 1 | No | No | Decreased and virally suppressed at 10wk postpartum | 0.56 to 0.64 (+14.2%) | 17 | 23 | **105*** | - |
|  |  |  |  |  | 16 | 21 | **193*** | - |
| 3 | No | Yes | Transient decrease, remained virally suppressed | 0.52 to 0.47  (-9.6%) | 20 | 17 | 27 | 18 |
|  |  |  |  |  | 14 | 19 | 26 | 22 |
| 4 | No | Yes | Transient decrease, remained virally suppressed | 0.57 to 0.69 (+21.1%) | 14 | 20 | 16 | 14 |
|  |  |  |  |  | 15 | 22 | 17 | 16 |
| 6 | No | Yes | Transient decrease, never reached viral suppression | 0.70 to 0.69  (-1.4%) | 16 | 22 | - | 33 |
|  |  |  |  |  | 16 | 27 | - | 83 |

** LFT above two times the upper limit of normal (>76 U/L for AST, >70 U/L for ALT).*

**Supplementary Figure 1:** A. Distribution of AST in women with high-risk HBV across study timepoints. B. Distribution of ALT in women with high-risk HBV across study timepoints.

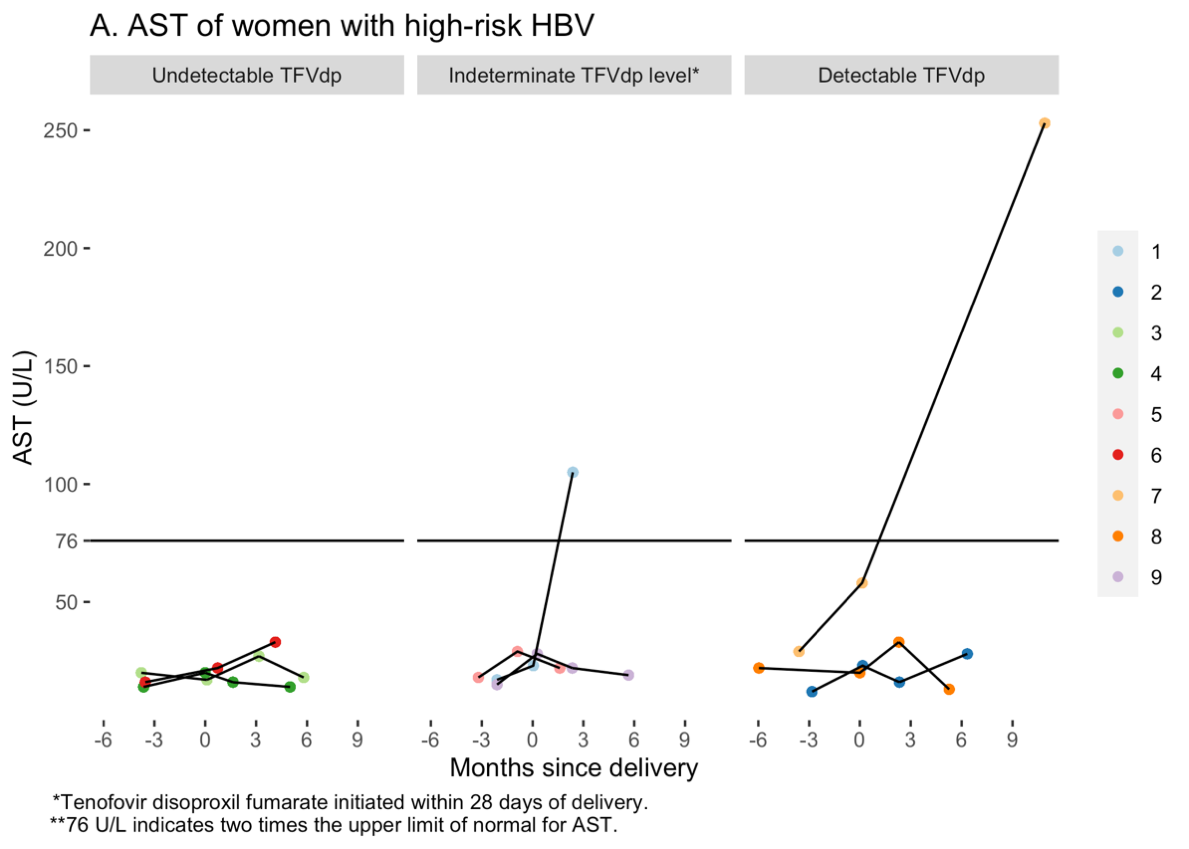

**A)**

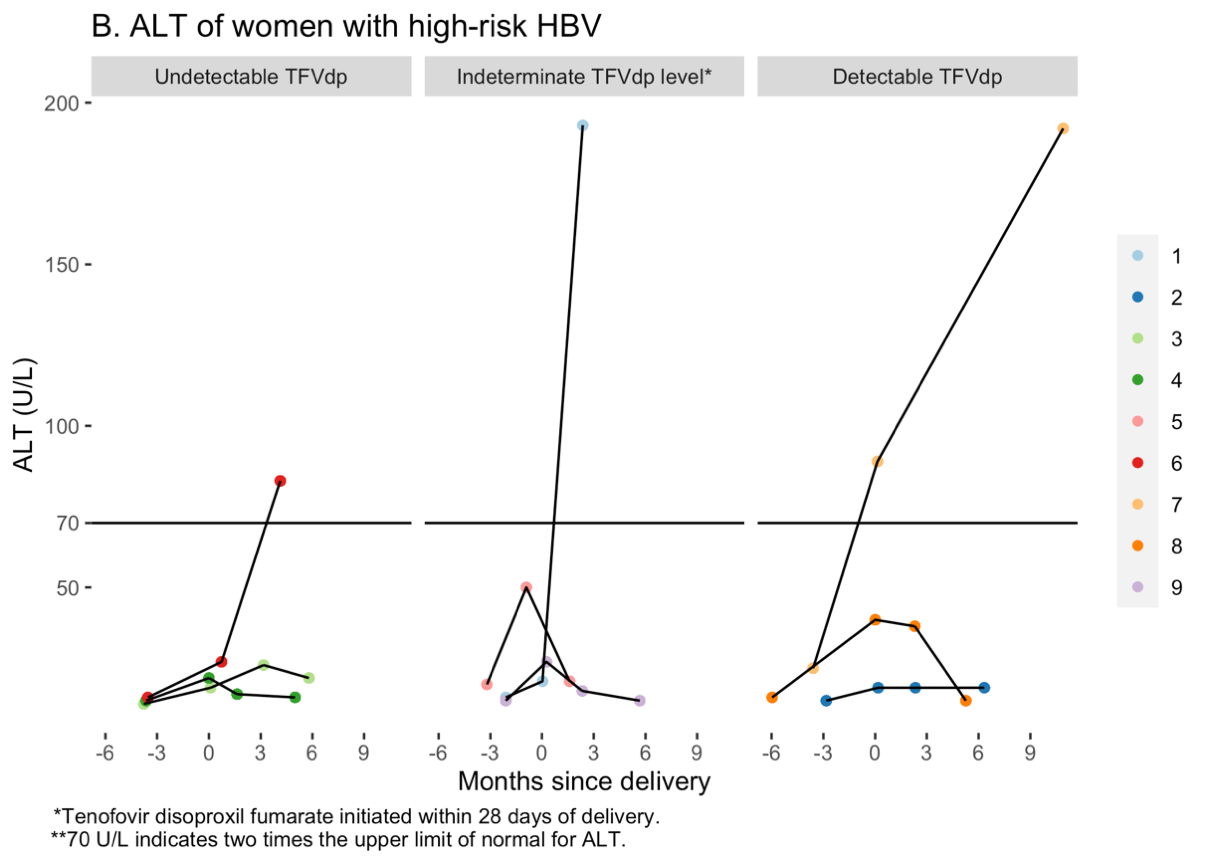
**B)**
